# Polygenic Risk and Genomic Signals in Premature Coronary Artery Disease

**DOI:** 10.64898/2026.10.23.26366138

**Authors:** Colin Laird, Diensn Xing, Mohammad Nahian Ferdous Abrar, Sambodh Sinha, Anderson Dewitt, Claire White, Kalgi Modi, Md Shenuarin Bhuiyan, Arif Yurdagul, Chris Kevil, A. Wayne Orr, Oren Rom, Steven A. Conrad, Mohammad Alfrad Nobel Bhuiyan

## Abstract

**Background:** Premature coronary artery disease (pCAD) is a distinct presentation of CAD. Using the All of Us (AoU) Research Program data, we compared pCAD with later-onset CAD across cardiometabolic, polygenic, and genome-wide domains.

**Methods:** We conducted a retrospective, cross-sectional case-case analysis of All of Us Research Program (AoU) participants with established CAD. pCAD was defined as first CAD event before age 55 years in males and before age 65 in females; later-onset CAD was defined as the first CAD event at or after age threshold. Pre-index cardiometabolic measures were extracted from electronic health records and modelled with logistic regression. Using 607,719 variants, a CAD polygenic risk score (PRS) was constructed using published summary statistics and short-read whole-genome sequencing data (srWGS). Results were standardized within self-reported race groups, and evaluated overall, by race, and with a PRS x race interaction. A case-case genome-wide association study (GWAS) compared pCAD with later-onset CAD.

**Results:** The CAD cohort included 49,919 participants, including 18,455 with pCAD and 31,464 with later-onset CAD. Participants with pCAD had higher BMI, LDL cholesterol and diastolic blood pressure, and lower HDL cholesterol, serum calcium, and systolic blood pressure. Higher BMI and LDL cholesterol were positively associated with higher odds of pCAD, whereas higher HDL cholesterol, systolic blood pressure, and serum calcium were inversely associated. PRS was associated with greater odds of pCAD in the genomic models (OR per 1-SD higher PRS, 1.14; 95% CI, 1.12–1.17; *P*<0.001). Analyses stratified by race showed PRS association was present among White participants but was not observed among Black participants. In the case-case GWAS, one variant reached genome-wide significance at 2q24.3 (rs13383835; OR=1.20; 95% CI, 1.13-1.27; *P*=5.8x10^-9^). A total of 137 variants met an exploratory threshold of *P*<1x10^-5^.

**Conclusions:** Premature onset of CAD is characterized by a distinct cardiometabolic profile driven by modifiable risk factors, greater polygenic burden, and genomic signals associated with earlier disease timing. A European-derived PRS did not transfer to Black participants, who were overrepresented in pCAD. Together, these findings support pCAD as a clinically and genetically distinct presentation of CAD and highlight the need for ancestry-specific validation of genomic risk tools in diverse cohorts.

## Introduction

Coronary artery disease (CAD) remains a leading cause of mortality in the United States and globally.^1^ Clinically, CAD is the pathological process in which the arteries supplying the heart become narrowed as a result of the accumulation of cholesterol plaques on the intimal surface of arterial vessels.^2^ In clinical practice and in most research, CAD is treated as a homogeneous disease, and age at onset is rarely used to distinguish disease phenotype.^3^ Yet there are a substantial number of patients who develop CAD prematurely – before age 55 in men and 65 in women.^3^ These patients at earlier risk have a longer timeline for which they are susceptible to recurrent cardiovascular events and have the most potential to benefit from early detection.^4^ Historically, these patients were the least likely to be identified: among adults younger than 50 presenting with their first myocardial infarction in the YOUNG-MI registry, only 49% would have met statin-eligibility criteria under prevailing guidelines.^5^

Fortunately, prevention guidelines are moving towards identifying risk in this population. The 2026 dyslipidemia guidelines adopted the PREVENT equations, extending the risk estimation to patients as young as 30 years old, making an estimated 21.5 million patients eligible for statin consideration.^6,7^ This shift is timely: between 1999 and 2024, deaths from later-onset CAD fell by 63%, whereas deaths from pCAD fell only 38%.^8^ A substantial fraction of CAD mortality continues to occur before the sex-specific premature threshold, even as overall CAD deaths are falling (*Figure 1*).

**Figure 1:**
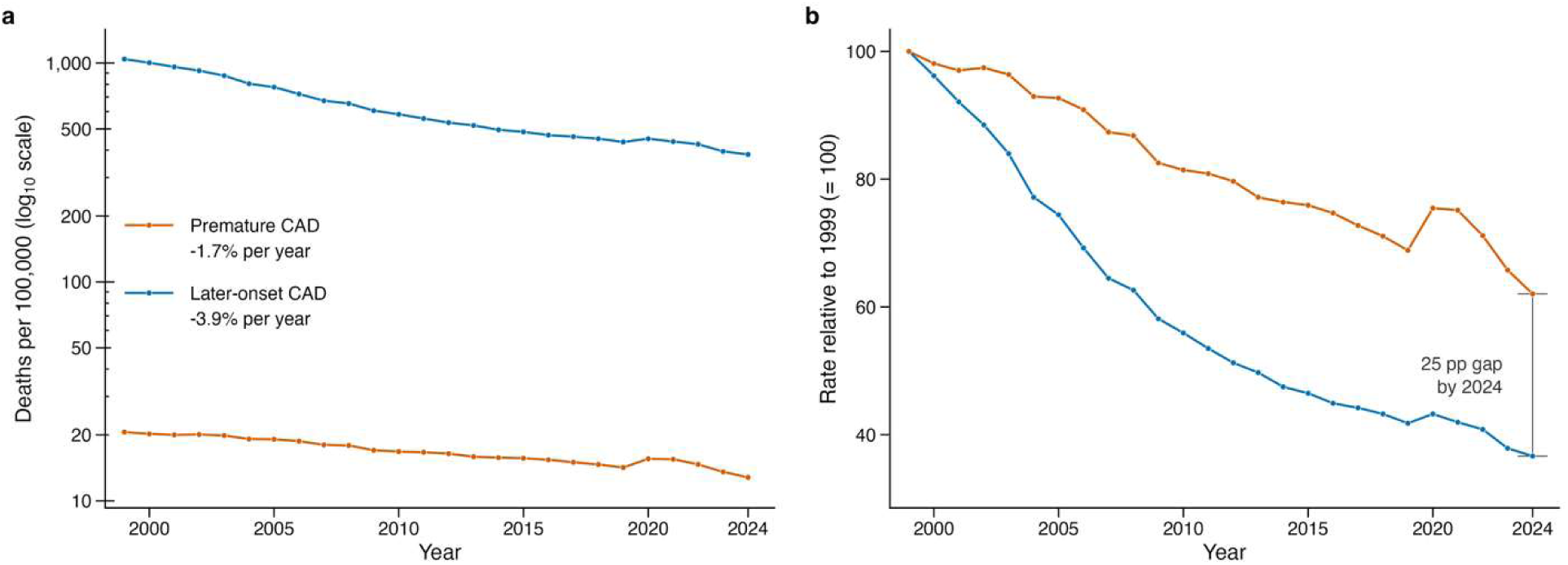
pCAD Mortality Trend. Premature coronary artery disease mortality has declined more slowly than later-onset disease in the United States. a) Crude death rates per 100,000 population on a logarithmic axis. The average annual percentage change across the period of 2019-2024 is shown. b) The same data was indexed to 1999, where the red line (pCAD) indicates a smaller relative decline. The bracket shows the difference in overall decline at 2024: 38% for pCAD vs. 63% for later-onset CAD.^8^

Whether pCAD represents a distinct disease phenotype remains to be answered. Well-established risk factors including hypertension, diabetes, obesity, metabolic dysfunction, smoking, and dyslipidemia contribute substantially to risk across age groups, and tools to predict cardiovascular events have improved substantially in the past two decades.^2,9^ To resolve this gap, we directly compared individuals with pCAD and later-onset CAD to isolate the factors that distinguish *early-onset* from *later-onset* disease. What distinguishes individuals who develop pCAD decades before other patients?

Genetic contributions play an important role in pCAD development, which is more heritable than later-onset CAD. Although both monogenic and polygenic factors contribute to pCAD risk, monogenic variants account for only a minority of pCAD incidence.^3^ The most prevalent monogenic contribution is familial hypercholesterolemia with the three most common mutations present in the low-density lipoprotein receptor (*LDLR*), apolipoprotein B (*APOB*), and proprotein convertase subtilisin/kexin 9 (*PCSK9*) genes.^3^ While monogenic contributions are well-established, the genetic architecture is predominately polygenic.

Polygenic risk scores (PRS) have emerged as a method of quantifying genetic risk by aggregating the effects across common variants of the human genome.^10^ Care must be taken in conveying what these scores represent, however. Though PRS are commonly communicated as “genetic predisposition” to a phenotype, a more appropriate description is that they are probabilistic estimates of risk based on populations’ genetic associations.^11^ Higher CAD polygenic risk has been associated with greater lifetime CAD risk and earlier disease onset in larger cohort studies.^12^ However, nearly all CAD PRS studies have been conducted with participants of European ancestry, limiting their generalizability to the diverse patient populations in the United States and raising the question of whether their use could widen health disparities.^13^ Because the populations in the U.S. that bear a disproportionate risk are also the least represented in genetic studies, evaluating these associations in a diverse cohort is important for determining whether these tools perform consistently across populations.^13^

We addressed these problems by analyzing three questions in the All of Us (AoU) Research Program. First, is cardiometabolic profile preceding pCAD different from the cardiometabolic profile preceding later-onset CAD? Second, does polygenic risk contribute to when CAD develops? Lastly, are there any genetic variants associated with pCAD that conventional case-control genetic association studies have not detected? (*Figure 2*)

**Figure 2:**
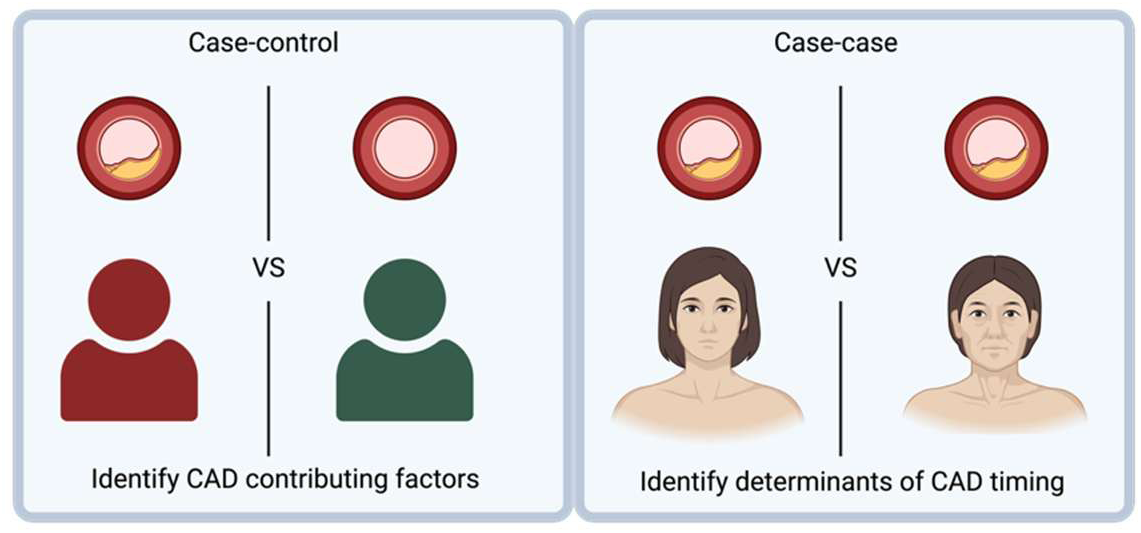
Conventional case-control compared individuals with CAD to unaffected/healthy individuals which identifies factors that influence whether disease develops at all while a case-case design allows for more effective targeting of the factors that affect disease timing.

## Methods

The AoU Research Program provides researchers with the unique opportunity to easily access and investigate complex disease phenotypes using linked electronic health record (EHR), demographic, and genomic data.^14^ The program’s protocol was approved by the All of Us Institutional Review Board, and all participants provided written informed consent. Our analysis of de-identified data was determined to be not human subjects research by the Louisiana State University Health Sciences Center Shreveport Institutional Review Board.

### Data Availability Statement

AoU data are available to registered researchers through the Researcher Workbench at researchallofus.org and cannot be exported or shared by the authors. GWAS summary statistics will be made available on Github. Analysis code is available upon request from M.A.N.B. C.P.L. had full access to all the data in the study and takes responsibility for its integrity and the data analysis.

### Analytic Cohort Definition Inclusion/Exclusion Criteria

Participants were enrolled in the AoU Research Program starting May 2018 with an enrollment age of adults aged 18 years and older from a network of over 340 enrollment sites across the United States.^14^ All analysis was performed in the researcher workbench and queried from Controlled Tier version 8. A CAD cohort was developed using a list of the relevant International Classification of Diseases (ICD) 9 and 10 codes, and from procedure records captured in ICD-9-Proc, HCPCS, CPT-4, and SNOMED concepts including coronary artery disease, myocardial infarction, ischemic heart disease, coronary atherosclerosis, percutaneous coronary intervention, coronary stent placement, angioplasty, or coronary artery bypass.^15^ A comprehensive list of diagnostic and procedure codes is available in *Tables S1 and S2*. A patient was included if they had one or more values for a CAD-defining diagnosis or procedure and an index date was established as the earliest CAD-related diagnosis or procedure date. Demographic variables included age, race, ethnicity, and sex at birth. Age was calculated by using difference between the birthdate of an individual and CAD index date. pCAD was defined as first CAD event before age 55 years in male participants and before age 65 years in female participants. Later-onset CAD was defined as a first qualifying CAD event at or after the corresponding sex-specific age threshold. Using this cohort definition, data was queried from EHR records that included demographics, condition codes, procedure codes, and relevant cardiometabolic measurements. Participants with missing demographic variables required for a given model were excluded from that analysis, and biomarker models were restricted to participants with available valid measurements for the corresponding cardiometabolic measure. For these analyses, participants were required to have at least one valid measurement before the CAD index date. After restriction to participants with recorded male or female sex at birth, the final analytic CAD cohort included 49,919 participants, including 18,455 with pCAD and 31,464 with later-onset CAD *(Figure S1*).

Genomic analyses were restricted to participants with available short-read whole-genome sequencing (srWGS) data and genetic principal components (*Figures S1 and S2*). Because genomic data availability and model covariate requirements differed by analysis, genomic sample sizes are reported separately for each model. The PRS analytic subset included 40,990 participants, of whom 15,004 had pCAD and 25,986 had later-onset CAD.

### Genome-Wide Association Study

We conducted a genome-wide association study (GWAS) to identify variants associated with pCAD compared to a cohort with later-CAD. The rationale for this design was to identify variants with premature disease presentation among CAD patients, as opposed to measuring genetic susceptibility to CAD in the general population. By comparing individuals with pCAD versus later-onset CAD, this design prioritizes variants associated with disease timing while reducing emphasis on variants associated with overall CAD susceptibility.^16^ Genomic analyses were performed using the All of Us Research Program Controlled Tier version 8 srWGS single nucleotide variant and insertion/deletion dataset. Genotype data were accessed using PLINK 2 (pgen) files from the All of Us WGS Allele Count/Allele Frequency (ACAF) threshold callset aligned to the Genome Reference Consortium Human Build 38 (GRCh38) reference genome.^17,18^ Association testing was conducted within the AoU researcher workbench. The genomic analysis cohort was derived from the analytic CAD cohort after filtering for genomic data and covariate availability. Participants were filtered and excluded from genomic analysis if they were younger than 18 years at first CAD event or if sex at birth was not recorded. The genomic analytic cohort was restricted to participants with CAD, available genomic data, valid CAD classification, and available genetic ancestry principal components (PC). The GWAS sample size included 38,427 participants: 13,798 with pCAD and 24,629 with later-onset CAD.

Sample-level quality control was applied before testing. Participants flagged for relatedness or sample-level genomic quality control concerns in the AoU auxiliary files were excluded. The first 10 genetic principal components were used from the All of Us ancestry prediction data and appended to the genomic dataset (*Figure S3*). For each chromosome, the ACAF pgen files were queried with PLINK 2. Variant quality control was applied within the analytic sample, and analysis was restricted to biallelic variants with minor allele frequency ≥ 0.01 and call rate ≥ 0.98. Association testing used PLINK 2 with logistic regression using an additive model. The GWAS model tested the association between each variant and odds of pCAD versus later-onset CAD, adjusting for sex and the first 10 genetic principal components to account for population ancestry structure. Age at onset was not included as a covariate because of the age-threshold case definition. Genome-wide significance was defined as *P*<5x10^-8^.^19^ Suggestive significance was defined as *P*<1x10^-5^.^20^ GWAS results were assessed using Manhattan and quantile-quantile (Q-Q) plots (*Figure S4*). Genomic inflation was evaluated using genomic control inflation factor (λ*_GC_*), which is calculated as the median observed chi-square statistic divided by the expected median (0.4549) of a chi-square distribution with 1 degree of freedom. This was then standardized to 1,000 cases and 1,000 controls (λ_1000_) given the cohort imbalance of cases and controls (later-onset CAD). ^21^ As a sensitivity analysis, the lead variant chi-square statistic was divided by λ*_GC_* to obtain corrected *P* value. Lead loci were defined by physical distance (250-kb windows) and annotated with Ensembl VEP v116 (Supplemental Methods).^22^

### CAD Polygenic Risk Score

A CAD polygenic risk score for each participant was constructed using a clumping-and-thresholding approach based on published GWAS summary statistics and AoU genomic data.^23^ The base GWAS was a large CAD meta-analysis by Aragam et al., which included more than one million participants and reported genome-wide association summary statistics for CAD.^24^ Because the base GWAS was derived largely from non-Hispanic White and East Asian participants, PRS results were interpreted with attention to potential limitations in transferability across ancestry groups.^13^ Base-data quality control, liftover from GRCh37 to GRCh38 using Hail and UCSC chain files, allele alignment, clumping parameters, and the P-value thresholds are described in the Supplemental Methods.^18,25,26^

PRS associations were evaluated across eight P-value thresholds (Supplemental Methods). The P≤1 score, which showed the strongest association with pCAD, was used as the primary PRS. Because the European-derived score’s distribution differed markedly across self-reported race groups, the PRS was standardized to mean 0 and SD 1 within each race group for all models in which it was the exposure, such that a 1-SD increment represents the same within-group quantity across every group. Logistic regression was used to estimate the association between the PRS and odds of pCAD. The primary model adjusted for sex and PC1–PC10, race-stratified models used the same covariates within each group, and effect modification was tested with a PRS × race interaction term. Results are reported as odds ratios (OR) and 95% confidence intervals for pCAD per 1-SD higher PRS. The association between standardized PRS and age at CAD presentation was visualized with scatterplot smoothing using LOESS and a 95% confidence band was created. *<u>Cardiometabolic and Laboratory Measurements:</u>* Clinical measurements were obtained from the EHR records linked to the analytical cohort using participant identification numbers and CAD index date. Measurements were restricted to values recorded before the CAD index date, defined previously as the earliest qualifying CAD-defining diagnosis or procedure date. Clinical and laboratory measures were queried from the All of Us measurement table using Logical Observation Identifiers Names and Codes (LOINC) concepts. The measurement panel included systolic blood pressure, diastolic blood pressure, body mass index, high-density lipoprotein cholesterol, low-density lipoprotein cholesterol and calcium. Participants were required to have at least one valid pre-index measurement for a given cardiometabolic measure. Pre-index values were averaged for each participant after appropriate unit conversion and exclusion of implausible values (Supplemental Methods).

### Statistical Analysis

Covariates were assessed for missingness, and each model was filtered to participants with complete data for the outcome, exposure, and covariates included for a particular model (*Table S3*). Sample size was reported separately for each analysis because availability differed across analytic categories. Prior to model fitting, diagnostics included evaluation of covariate distribution and phenotype counts across demographic and genetic principal components. Statistical analysis was performed using Python using statsmodels for regression modeling.^27^ Statistical significance was evaluated using two-sided *P* values from the corresponding regression models. For simplicity, “clinical” models refer to the cohort adjusted for demographics (race, sex, and ethnicity) and “genomic” models refer to the genotyped cohort adjusted for sex, PRS, and PC1-PC10 to assess whether cardiometabolic measurements are independent of these variables. EHR-derived measurements, risk scores, and genetic principal components were not uniformly available across participants, so analytic sample size varied across models according to model-specific variable availability (*Table S4*).

Univariable and multivariable logistic regression models were used to estimate associations between cardiometabolic measures, PRS, and odds of pCAD versus later-onset CAD. The primary outcome for regression models was pCAD status. For each primary predictor, an unadjusted model was first fit to estimate the association with the odds of pCAD. Race and ethnicity were self-reported in the All of Us participant survey. Race was categorized as White, Black or African American, Asian, American Indian or Alaska Native, more than one population, other (Middle Eastern or North African, Native Hawaiian or Other Pacific Islander, or none of these), or unknown (skipped or preferred not to answer); ethnicity was categorized as Hispanic or Latino, not Hispanic or Latino, or unknown. Continuous exposures and risk scores were standardized prior to performing logistic regression. Standardized variables were modeled per 1-standard deviation higher value. For each model, odds ratios and 95% confidence intervals were calculated. The primary model was adjusted for sex, race, and ethnicity. Modeling with risk scores included adjustment for the first 10 genetic principal components to reduce population structure confounding. The primary risk model was:

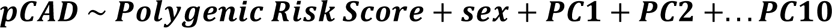

Each cardiometabolic measure was separately modeled as a continuous predictor. Additional models included adjustment for genetic principal components when evaluating genomic predictors or models incorporating PRS. Unadjusted models were also included for comparison with multivariable models.

## Results

### Case-Case GWAS of pCAD

The GWAS included 38,427 participants with CAD (13,798 with pCAD and 24,629 with later-onset CAD). Mean age at CAD index date was 51.4 years among participants with pCAD and 69.7 years among participants with later-onset CAD. Among 6,934,805 variant associations, the genomic inflation factor was λ_GC_=1.12 (λ_1000_=1.007) (*Figure 3 and Figure S4*). An intronic variant at 2q24.3 within a long non-coding RNA (rs13383835; OR 1.20; 95% CI, 1.13–1.27; *P*=5.8x10^-9^) reached genome-wide significance. After genomic-control correction, rs13383835 remained significant (*P*=4.0×10⁻⁸). A total of 137 variants met an exploratory threshold of *P*<1x10^-5^. The strongest suggestive protein-coding candidates were intronic variants in the *THRB* region (rs6798561; OR 1.50; 95% CI, 1.29–1.76; *P* = 2.2x10^-7^), *SORCS3* region (rs2027113; OR 1.10; 95% CI, 1.06–1.15; *P* = 7.6x10^-7^), and *AHR* region (rs73680639; OR 0.85; 95% CI, 0.79–0.90; *P* = 6.3x10^-7^). Annotated lead loci are reported in *Table S5*.

**Figure 3:**
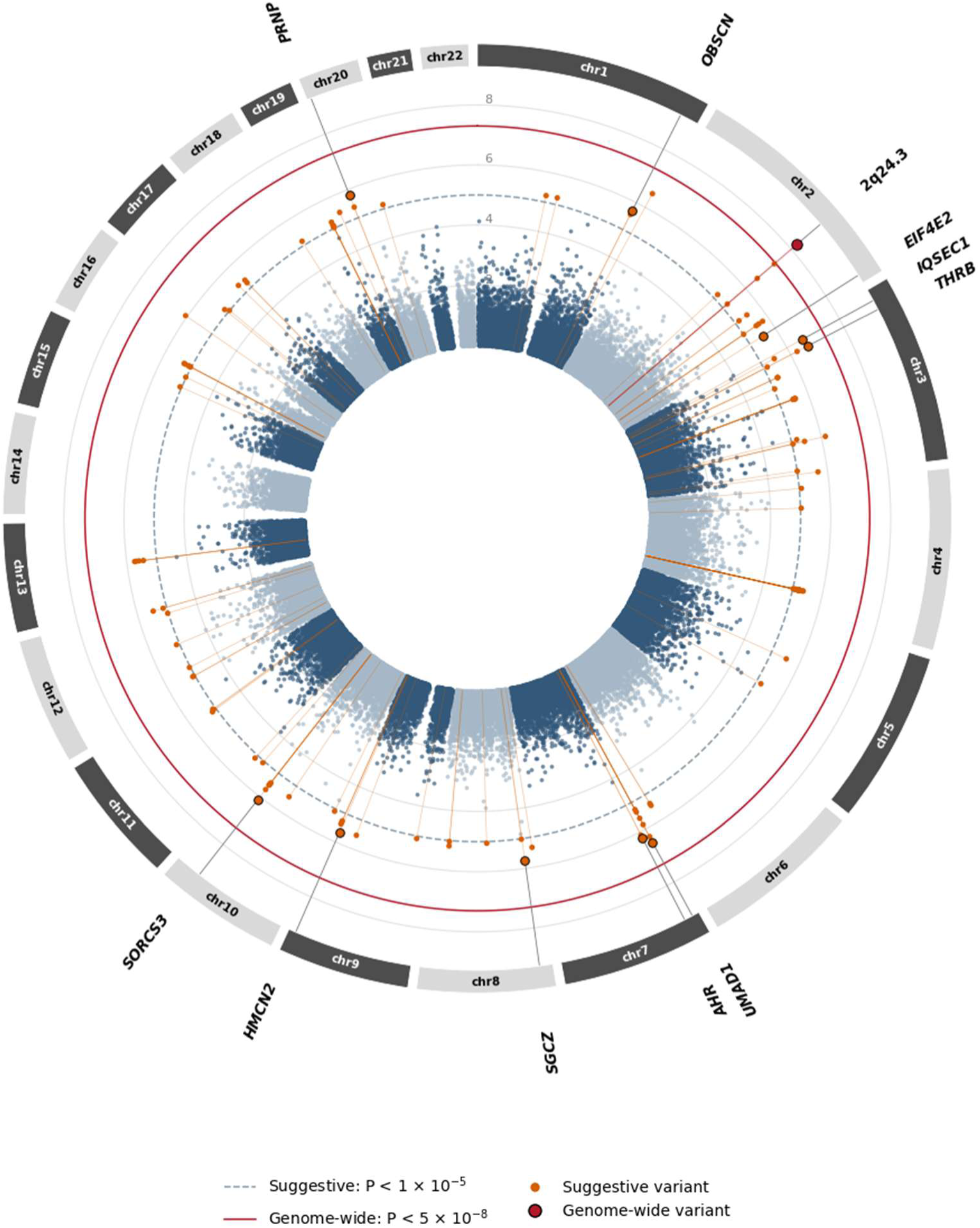
Circular Manhattan plot of the case-case genome-wide association study for premature versus later-onset CAD. Each point represents a genetic variant. Distance from the inner rings corresponds to −log_10_(P) for the association with premature versus later-onset CAD (n = 38,427; genomic control λ_GC_ = 1.12). The dashed circle marks the suggestive threshold (P<1x10^-5^) and the solid circle the genome-wide threshold (P<5x10^-8^). The genome-wide significant variant at 2q24.3 is shown in dark red. Suggestive loci are labeled with their nearest gene.

Demographic characteristics differed between pCAD and later-onset CAD. Female participants represented 67.6% of the pCAD group and 31.9% of the later-onset CAD group. Race and ethnicity also differed by pCAD status. Black participants made up 24.7% of the pCAD group and 11.1% of the later-onset CAD group. Hispanic or Latino participants represented 18.1% of the pCAD group and 9.8% of the later-onset CAD group. White participants represented a larger proportion of the later-onset CAD group (70.7%) (*Table 1*).

**Table 1:** Baseline characteristics of patients with premature versus later-onset coronary artery disease. Race and ethnicity were self-reported. Other includes American Indian or Alaska Native (reported in Table S8), Middle Eastern or North African, Native Hawaiian or Other Pacific Islander, and none of these. Unknown includes skipped, prefer not to answer, and not indicated. † P<0.001 (Mann–Whitney U test for continuous variables and chi-square test for categorical variables). BMI indicates body mass index; HDL, high-density lipoprotein; LDL, low-density lipoprotein; SD, standard deviation.

| Characteristic | pCAD (n=18,455) | Later-onset CAD (n=31,464) | P-value |
| --- | --- | --- | --- |
| **Demographics** |  |  |  |
| Age at CAD index, mean ± SD | 51.30 ± 9.04 (n=18,455) | 69.76 ± 7.63 (n=31,464) | <0.001<sup>†</sup> |
| Sex, n (%) |  |  | † |
| Female sex | 12,479 (67.6%) | 10,034 (31.9%) |  |
| Male | 5,976 (32.4%) | 21,430 (68.1%) |  |
| Race, n (%) |  |  | † |
| White | 8,636 (46.8%) | 22,259 (70.7%) |  |
| Black or African American | 4,556 (24.7%) | 3,490 (11.1%) |  |
| Asian | 290 (1.6%) | 472 (1.5%) |  |
| More than one population | 1,008 (5.5%) | 1,036 (3.3%) |  |
| Other | 744 (4.0%) | 838 (2.7%) |  |
| Unknown | 3,221 (17.5%) | 3,369 (10.7%) |  |
| Ethnicity, n (%) |  |  | † |
| Hispanic or Latino | 3,348 (18.1%) | 3,079 (9.8%) |  |
| Not Hispanic or Latino | 14,555 (78.9%) | 27,290 (86.7%) |  |
| Unknown | 552 (3.0%) | 1,095 (3.5%) |  |
| **Primary Cardiometabolic measures** |  |  |  |
| Body Mass Index, kg/m<sup>2</sup> | 32.85 ± 8.88 (n=12,234) | 29.88 ± 6.34 (n=20,758) | |
| Systolic blood pressure, mmHg | 129.98 ± 16.07 (n=11,623) | 131.94 ± 13.73 (n=22,410) | |
| Diastolic blood pressure, mmHg | 78.70 ± 9.89 (n=11,622) | 75.67 ± 8.28 (n=22,395) | |
| HDL cholesterol, mg/dL | 49.07 ± 16.05 (n=9,523) | 50.41 ± 15.63 (n=17,861) | |
| LDL cholesterol, mg/dL | 112.32 ± 38.24 (n=1,902) | 102.10 ± 32.45 (n=3,956) | |
| Serum calcium, mg/dL | 9.11 ± 0.49 (n=13,632) | 9.21 ± 0.46 (n=23,693) | |

### Cardiometabolic Profiles

Cardiometabolic profiles differed between groups. Participants with pCAD had higher BMI than participants with later-onset CAD (32.85 ± 8.88 vs 29.88 ± 6.34 kg/m^2^), higher DBP (78.70 ± 9.89 vs 75.67 ± 8.28 mmHg), and higher LDL cholesterol among participants with available LDL measurements (112.32 ± 38.24 vs 102.10 ± 32.45 mg/dL). Participants with pCAD also had lower SBP (129.98 ± 16.07 vs 131.94 ± 13.73 mmHg), lower HDL cholesterol (49.07 ± 16.05 vs 50.41 ± 15.63 mg/dL), and lower serum calcium (9.11 ± 0.49 vs 9.21 ± 0.46 mg/dL). All descriptive comparisons were statistically significant at *P*<0.001 (*Table 1*). In modeling adjusted for sex, PRS, and PC1–10, each 1-SD higher BMI (OR 1.36; 1.33–1.40) and diastolic blood pressure (OR 1.53; 1.49–1.58) was associated with higher odds of pCAD, whereas higher systolic blood pressure (OR 0.80; 0.78–0.82), HDL cholesterol (OR 0.70; 0.67– 0.72), and serum calcium (OR 0.79; 0.77–0.81) were associated with lower odds (*Table S6 and Figure S5A*). LDL cholesterol, available in a smaller subset, was associated with higher odds of pCAD in both the demographic-adjusted (OR 1.26; 1.18–1.34; n=5,858) and genomic (OR 1.25; 1.17–1.34; n=4,806) models (*Table S7 and Figure S5A*).

### Polygenic Risk

Among White participants, standardized CAD PRS was highest among those presenting at the youngest ages and declined steadily with older age at presentation (*Figure 4*). PRS was above the mean among earlier-presenting participants and progressively lower among those presenting at older ages, corresponding to a slope of −0.011 SD per year of age (*P*=6x10^-92^). Each 1-SD higher PRS was associated with higher odds of pCAD after adjustment for sex and PC1–10 (OR 1.14; 95% CI, 1.12–1.17; P<0.001; *Table S6*). The association differed by race: OR 1.22 (95% CI, 1.18–1.26) among White, 1.00 (0.94–1.06) among Black, and 1.57 (1.10–2.24) among Asian participants (Tables S8 and S9 and Figure S5B).

**Figure 4:**
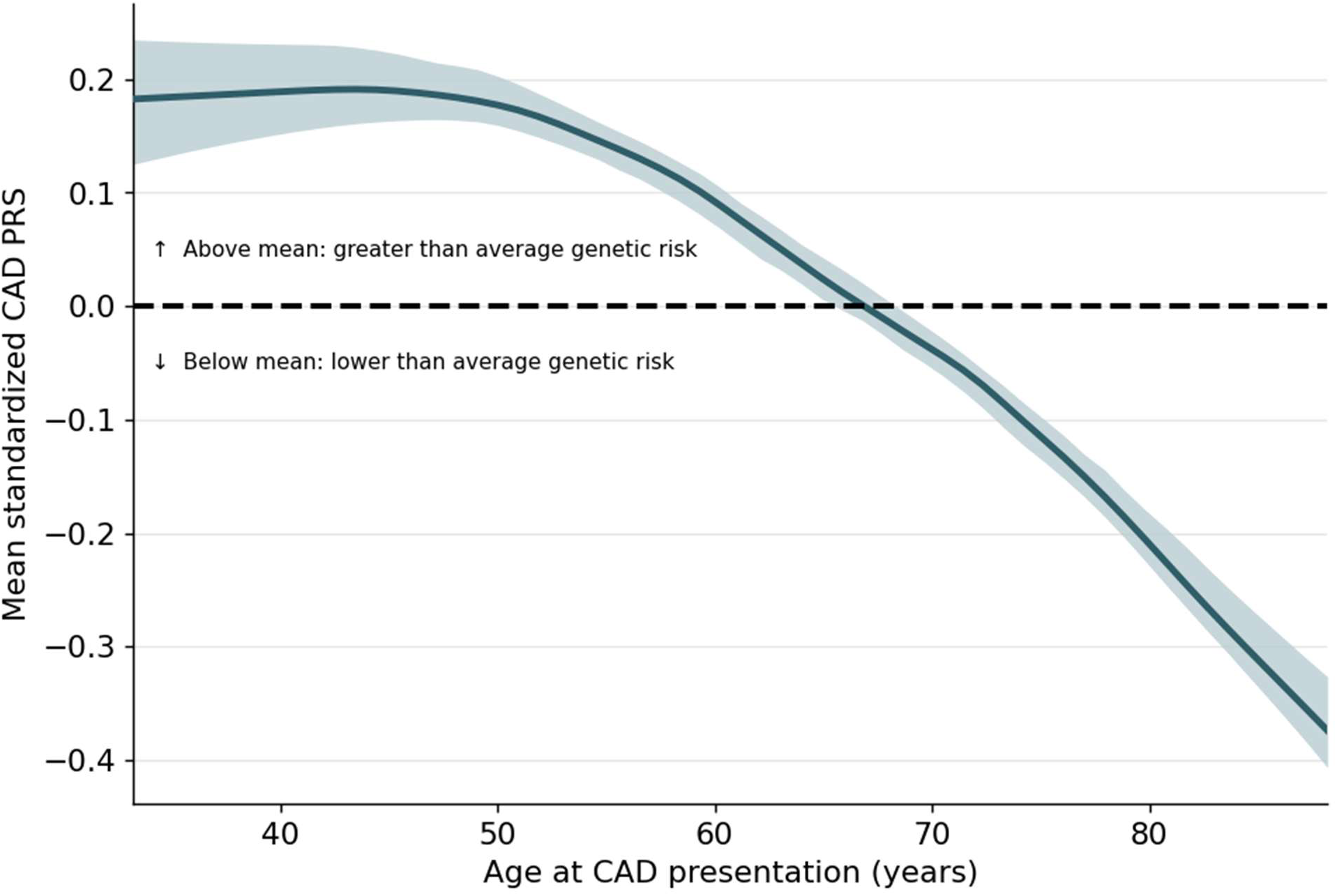
Coronary artery disease polygenic risk score decreases with older age at presentation. Mean standardized CAD polygenic risk score as a function of age at CAD presentation among White participants (n = 25,665). The shaded region is a 95% confidence band interval.

## Discussion

In a cohort of nearly 50,000 participants with CAD, pCAD differed across three dimensions: lab measurements, higher PRS associated with earlier disease and uneven distribution of risk across ancestry groups, and genomic signals associated with earlier disease presentation. These findings suggest that earlier disease is phenotypically distinct from later CAD and demonstrates that European genomic data does not adequately perform across the diverse U.S. population if used to calculate polygenic risk scores.

### Cardiometabolic Profile of pCAD

Participants with pCAD had higher BMI, diastolic blood pressure, LDL, and lower systolic blood pressure, HDL, and serum calcium. The pulse pressure association observed in this study is consistent with blood pressure physiology that changes as age progresses. Pulse pressure widens with older age as arterial stiffening increases, systolic pressure rises, and diastolic pressure declines after midlife.^28^ The pCAD cohort in this study had a mean age of roughly two decades younger so this observed pulse pressure gradient is more consistent with age-related arterial stiffening than early disease pathology. The BMI, LDL, and HDL differences, however, are more consistent with modifiable risk factors instigating pCAD and less related to age related physical decline, reiterating the well-understood patterns for premature heart disease.^29^ We observed lower serum calcium among pCAD patients. Prospective cohort studies have linked higher circulating serum calcium to increased risk of CAD.^30^ However, these studies compared individuals who develop CAD versus those who did not, whereas this study compares the measurements among the *timing* of the disease. Even so, the absolute difference was minimal (9.11 mg/dL vs 9.21 mg/dL) and likely not enough to implicate calcium in a direct role for pCAD.

### PRS Association

CAD polygenic risk was independently associated with 14% greater odds of pCAD after adjusting for sex and principal components, supporting a genetic contribution to earlier disease presentation. Among White participants, PRS steadily declined with older age at first disease presentation. A prior study showed that CAD PRS had prognostic value in predicting myocardial infarction in younger adults, and our analysis builds on that foundation to show that among people who develop CAD, higher polygenic risk is correlated with earlier onset.^31^ These findings support our hypothesis that pCAD is distinct from later-onset CAD, and that it is driven by high genetic burden.

### GWAS

The data on case-case GWAS designs for CAD are sparse compared to the standard case-control design in which the genetic loci from a non-diseased phenotype are compared with the genetic variants of a population with a disease of interest, although a small number of GWAS have used within-disease stratification.^32,33^ The rationale for this design was to prioritize variants associated with disease timing among individuals who develop CAD, reducing the emphasis on well-studied variants driving overall CAD burden.^16^ Our GWAS identified one genome-wide significant locus in the 2q24.3 region, located within a long non-coding RNA with no proximal protein-coding gene. Genomic inflation (λ*_GC_*=1.12) in a multi-ancestry group may reflect polygenicity or population stratification. The λ*_1000_* was 1.007 and the lead variant remained significant after genomic control, but the 2q24.3 association requires replication in an independent sample. Among suggestive loci, intronic candidates included *AHR*, which mediates vascular responses to environmental exposures including tobacco smoke; *THRB*, encoding thyroid hormone receptor beta with established cardiometabolic effects; and *SORCS3*, a sortilin-family receptor related to the established *SORT1* CAD locus.^34–36^ These exploratory signals may reflect genetic differences associated with earlier CAD presentation, and the suggestive variants may reach genome-wide significance in larger cohorts.^37^ Because this GWAS is case-case in design, findings were interpreted as variants associated with earlier disease presentation among individuals with CAD rather than variants associated with CAD susceptibility in the general population. Within the scope of this study, these signals cannot be interpreted in terms of identifying causal genes. Many variants associated with complex phenotypes are non-coding and target regulatory mechanisms for genes in further regions.^36^ These loci are therefore best interpreted for replication in other cohorts and candidates for fine-mapping analysis.

We found genetic risk was not shared across ancestral groups. When split by subgroup, the PRS association in White participants had OR=1.22 but was 1.00 among Black participants. This was an expected consequence of deriving scores using base data from a European cohort and is consistent with literature that documents reduced PRS performance in individuals with African ancestry.^13^ This result coupled with the scarcity of diverse base data highlights the need for greater emphasis by researchers to produce ancestry-specific base data to facilitate studies that can more accurately calculate genetic risk in diverse cohorts.^13^ In our cohort, Black participants made up 24.7% of the pCAD group, but just 11.1% of the later-onset CAD group, a disproportionate representation that is consistent with higher burden and earlier onset of cardiovascular disease among Black Americans.^38^

### Limitations

This study is not without limitations. Race and ethnicity were self-reported and correlate imperfectly with genetic ancestry. PRS were derived from a predominantly European-ancestry base GWAS. Because the genome-wide analysis utilized regression with 10 principal components in a multi-ancestry sample, residual stratification cannot be excluded. The 2q24.3 locus still requires replication in an independent cohort. Additionally, factors such as smoking, medication use, and family history, may confound the cardiometabolic associations we found.

### Conclusion

Using a diverse cohort, we present a case-case analysis of participants with established CAD where pCAD was associated with a distinct cardiometabolic profile, including higher BMI, LDL cholesterol, and diastolic blood pressure, and lower HDL cholesterol, serum calcium, and systolic blood pressure when compared with later-onset CAD. These findings reinforce the observation that many patients developing pCAD would not be identified through age-based screening and risk calculation tools and support the development of earlier screening that is rooted in clinical measurements and inherited genetic risk. Polygenic risk scores were independently associated with greater odds of pCAD, but the association was not observed among Black participants. An exploratory case-case GWAS identified one genome-wide significant locus at 2q24.3 and multiple suggestive loci. Together, these findings support pCAD as a clinically distinct presentation of CAD with genetic differences and highlight the need for ancestry-specific validation of genomic risk tools in diverse cohorts.

## Acknowledgements

We gratefully acknowledge *All of Us* participants for their contributions, without whom this research would not have been possible. We also thank the National Institutes of Health’s *All of Us* Research Program for making available the participant data examined in this study.

## Supplemental Material

Supplemental Methods Tables S1-S9

Figures S1-S5

## Contribution Statement

This project was conceived by C.P.L and M.A.N.B., C.P.L assembled and analyzed the data, C.P.L., M.A.N.B., and A.W.O contributed to data interpretation. C.P.L drafted the manuscript. All authors read and contributed edits and approved the final manuscript.

## Non-standard Abbreviations and Acronyms

ACAF: allele count/allele frequency
AoU: All of Us Research Program
CAD: coronary artery disease
CPT-4: Current Procedural Terminology, 4th edition
DBP: diastolic blood pressure
EHR: electronic health record
GRCh38: Genome Reference Consortium Human Build 38
GWAS: genome-wide association study
HCPCS: Healthcare Common Procedure Coding System
ICD: International Classification of Diseases
λ_GC_: genomic control inflation factor
λ_1000_: genomic inflation factor standardized to 1,000 cases and 1,000 controls
LOESS: locally estimated scatterplot smoothing
LOINC: Logical Observation Identifiers Names and Codes
PC: genetic principal component
pCAD: premature coronary artery disease
PREVENT: Predicting Risk of Cardiovascular Disease Events (equations)
PRS: polygenic risk score
Q-Q: quantile-quantile
SBP: systolic blood pressure
SNOMED: Systematized Nomenclature of Medicine
SNP: single nucleotide polymorphism
srWGS: short-read whole-genome sequencing
VEP: Variant Effect Predictor (Ensembl)

